# Longitudinal Case-Control Study of Active and Passive Dense Mammographic Breast Tissue

**DOI:** 10.1101/2024.02.17.24302978

**Authors:** Kendra Batchelder, Basel White, Christina Cinelli, Amy Harrow, Christine Lary, Andre Khalil

## Abstract

Mammography is used as secondary prevention for breast cancer. Computer-aided detection and image-based short-term risk estimation were developed to improve the accuracy of mammography. However, most approaches inherently lack the ability to connect observations at the mammography level to observations of cancer onset and progression seen at a smaller scale, which can occur years before imageable cancer and lead to primary prevention. The Hurst exponent (*H*) can quantify mammographic tissue into regions of dense tissue undergoing active restructuring and regions that remain passive, with amounts of active and passive dense tissue that differ between cancer and controls at diagnosis. A longitudinal retrospective case-control study was conducted to test the hypothesis that differences can be detected before diagnosis and changes could signal developing cancer. Mammograms and reports were collected from 50 patients from Maine Medical Center in 2015 with at least a 5-year screening history. Age-matching patients within 2 years created a primary dataset, and within 5 years, a secondary dataset was created to test for sensitivity. The amount of passive (*H* ≥ 0.55) and active dense tissue (0.45 < *H* < 0.55) was calculated for each breast and was predicted by creating a linear mixed-effects model. Cancer status was a predictor for passive (*p* = 0.036) and active (*p* = 0.025) dense tissue using the primary dataset. However, when increasing the power, cancer status was a predictor for active dense tissue (*p* = 0.013), while breast status (*p* = 0.004), time (*p* = 0.009), and interaction (*p* = 0.038) were predictors for passive dense tissue. This suggests active dense tissue is a risk for cancer and passive dense tissue is an indication of developing cancer.

**Required Key Messages:**

- Mammographic dense breast tissue can be separated into regions of active and passive.
- There is more active dense breast tissue in pathology-confirmed cancer cases than controls.
- Increases in passive dense tissue in a breast could indicate a developing tumor.

## INTRODUCTION

Breast cancer stands as the most diagnosed cancer globally and is the second leading cause of cancer-related fatalities among women [1]. Addressing the high incidence rates of this disease through preventive measures can enhance patient outcomes and alleviate the burden of breast cancer on both public health and the economy. Cancer prevention is achieved through interventions, categorized as primary, secondary, and tertiary [2]. Considerable research has been devoted to secondary prevention strategies for breast cancer, aimed at advancing early detection, diagnosis, and removal of cancer and pre-cancerous conditions before they progress beyond their initial site through screening when treatment is most likely successful [3]. Screening guidelines are provided by entities such as the US Preventive Task Force [4], the American Cancer Society Field [5], the American College of Obstetrics & Gynecology [6], and the American College of Radiology [5, 6]. These guidelines recommend mammography, the only modality shown to decrease mortality [7], as the imaging modality for most women. Nevertheless, the extent of mortality reduction attributed to mammography screening ranges from 19% to 40%, contingent on age and breast density [7], with sensitivity varying from 86% to 89% in women with minimal dense breast tissue to 62-68% in those with highly dense breasts [8].

Recently, potentially modifiable risk factors have been causally linked to a wide range of cancers [9], and approximately 40% of cancers can be prevented by reducing risk factors and implementing primary prevention strategies [10]. Taken with the continued increase in incidence rates and with breast cancer becoming more common among younger women [11, 12], there is a growing emphasis on the primary prevention of breast cancer to hinder the start of the carcinogenic process. Risk models and genetic testing can help identify individuals at an increased risk of developing breast cancer [13]. However, known genetic predisposition or heredity plays a limited role in cancer, accounting for only 5% to 10% of all cancer cases [10]. Traditional risk models, such as the Tyrer-Cuzick, Gail, and Breast Cancer Surveillance Consortium (BCSC) models, are based on varying familial and personal health histories and some models are not calibrated for all populations [14].

Breast density has recently been recognized as one of the strongest independent risk factors for breast cancer, with women with dense breasts having a higher risk of developing breast cancer than women with non-dense breasts [15]. Incorporating breast density measurements has marginally improved some models’ predictive performance to ∼70% [16]. However, the association between breast density and its link to cancer remains unclear. In addition, the World Health Organization estimates that 50% of breast cancer cases do not have known identifiable risk factors [17], which creates a missed opportunity to provide enhanced surveillance or risk reduction methods to women at elevated risk to reduce both the societal and economic impact of breast cancer.

New efforts involve applying artificial intelligence to screening mammography to overcome the limitations of traditional approaches to breast cancer risk assessments. Several models that estimate breast cancer risk scores have been developed, including *Mirai, Globally-Aware Multiple Instance Classifier, MammoScreen, ProFound AI*, and *Mia*, and these models have better predictive performance at 0 to 5 years than the BCSC risk model that includes traditional risk factors (BCSC area under the receiver operator curve (AUC) = 0.61, AI algorithms’ AUCs= 0.63-0.67) [18]. Furthermore, advancements in radiomics have allowed for improved quantification and inclusion of parenchymal textural complexity and patterns into models to improve risk estimation beyond breast density [19]. However, AI approaches are not always generalizable to new settings and populations, such as races, ethnicities, and mammography equipment outside of the training set [20]. Furthermore, their generalizability has yet to be robustly demonstrated, with one study showing recall rates increased by 3-fold following mammography equipment software upgrades [20].

In addition, AI’s inherent lack of explainability and inability to link to known cancer dynamics plays a role in the hesitancy to adopt it in a clinical setting. The biophysical processes of tumor onset have been studied extensively at the cellular level [21-24]. Still, limited research has been done to explore which, if any, of these processes can lead to large-scale features that could be captured on screening mammograms. The development and progression of malignant tumors are intricately influenced by the cancer cells and the surrounding tissues and cells collectively known as the tumor microenvironment [7]. Comprising stromal cells, immune cells, extracellular matrix, and blood vessels, the tumor microenvironment interacts with cancer cells, crucial in promoting or inhibiting tumor growth and invasion. In breast cancer, the tumor microenvironment assumes particular significance, as events during breast development and exposure to various risk factors can reshape the breast microenvironment, establishing a permissive setting for cancer initiation and progression. It has been established that tumor onset and progression lead to disorganization and begin approximately 8 years before an imageable tumor [25]. Changes in breast tissue seen in mammography, including increased mammographic breast density, may be associated with elevated collagen levels and the structural organization of stroma, which influences tumor invasion dynamics [11, 14]. Therefore, a metric that can quantify subtle signs of dense breast tissue that is undergoing active restructuring vs passive dense breast tissue could provide further insights into developing abnormalities and the associated risk for breast cancer.

The 2D Wavelet-Transform Modulus Maxima (WTMM) method has been used in several fields to analyze complex signals to extract features and quantify spatial structure to gain insights into the underlying mechanisms of complex organizations [26-31]. In previous studies, the 2D WTMM method was employed to capture the structural organization of mammographic tissue, via the Hurst exponent (*H*), and the calculated organization was inferred to be linked to the structure of the tumor microenvironment at the time of diagnosis [32, 33]. The method allows for segmenting dense breast tissue into regions of active dense tissue, i.e., regions that show structural reorganization occurring and are inferred to be linked to the dynamics of cancer onset and progression, and regions of passive dense tissue. This research aims to computationally quantify mammographic breast tissue composition by detecting active and passive dense tissue regions and assess if longitudinal changes in the tissue differ between cancer cases and controls.

## METHODS

This study received IRB Approval with Waiver of Informed Consent/Authorization (IRB #4664) from Maine Medical Center (Portland, ME) on September 6, 2015, and was compliant with the Health Insurance Portability and Accountability Act (HIPAA).

### Cohort Description

“FOR PRESENTATION” mammographic images of the standard bilateral mammographic views, i.e. right and left mediolateral oblique (MLO) and cranial caudal (CC), from full-field digital mammography were retrospectively collected from Maine Medical Center (Portland, ME, USA) in 2015 from women with at least a 5-year screening exam history. Screen-detected breast cancer cases were confirmed to be malignant by biopsy within 12 months of the last screening exam. Controls had no history of cancer or benign breast disease. The tumorous breast, i.e. the breast that contained the pathology-confirmed malignancy, and the contralateral breast were identified in the accompanying pathology reports for the malignant cases. Breast density scores of A: almost entirely fatty, B: scattered areas of fibroglandular density, C: heterogeneously dense, or D: extremely dense, were assigned to mammogram exams by two expert breast radiologists (AH and CC) following the BI-RADS 5^th^ edition [34].

The primary dataset was created by age-matching patients using their age at the time of the last screening before diagnosis for malignant cases and the time of the last visit for controls. Using nearest neighbor logistic regression propensity score matching, eligible matches were restricted to be within 2 years of each other. Up to two controls were matched to each malignant case using the *MatchIt* function in R [35]. To test sensitivity and explore the outcomes associated with increasing the power, a second dataset was created with eligible matches being restricted within 5 years of each other.

### Analysis of Mammographic Images (Figure 1)

The analysis used the four standard bilateral mammographic views: right MLO, left MLO, right CC, and left CC. As a preprocessing step, black and white binary masks were generated through visual inspection. The breast tissue was contoured manually using the polygon feature in Fiji [36] to eliminate the image background, label, and pectoral muscle, and a mask that segmented the breast tissue was produced, which was then utilized for subsequent analysis (**Fig 1A-C**). A 360×360 pixel sliding window was positioned at the top left of the segmented breast tissue. The sliding window shifted from left to right and top to bottom with a step size of 32 pixels between subregions. If the central 256×256 of each subregion was entirely contained inside the mask, the subregion was accepted for further analysis (**Fig. 1D-H**). Each subimage was wavelet transformed across 50 different size scales. The corresponding maxima chains and their maxima, maxima lines, partition functions, *h*(*a, q*) and *D*(*a, q*) were generated following the methods described by Marin et al. [15] and Gerasimova-Chechkina [16]. Following these calculations only the central 256×256 pixels of each subimage was kept to mitigate edge effects (**Fig. 1I-K**).

**Figure 1:**
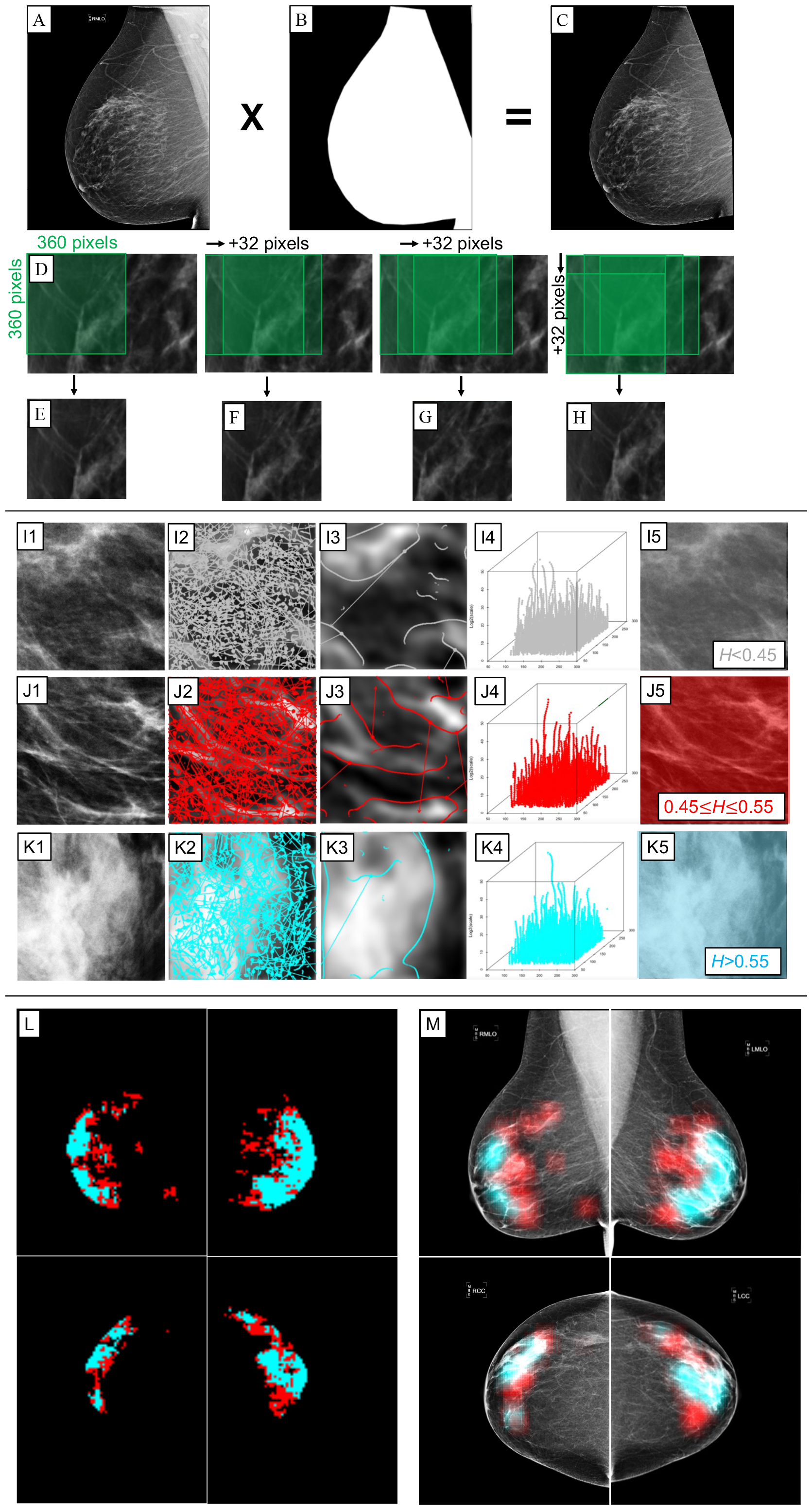
**Overview of the 2D WTMM multifractal sliding window approach** (Marin et al. [15] and Gerasimova-Chechkina [16]). Sliding window approach to divide mammographic images into subregions. A mammographic image (A) is used to create a mask (B). The mask is used to segment the breast tissue (C). A 360 pixels by 360 pixels box (D) is placed in the upper left corner of the image. The box is then moved by a step size of 32 pixels horizontally and vertically. If the box contained breast tissue and no background, the 360 pixels x 360 pixels subimage is kept for the analysis (E-H). To identify sub-types of mammographic breast tissue, subimages containing fatty tissue (I1), active dense tissue (J1) and passive dense tissue (K1) were wavelet transformed at 50 different size scales with scale a = 10 (I2, J2, K2) and scale a = 30 (I3, J3, K3) shown with the corresponding WTMM and WTMMM. The WTMMM were used to construct the WT skeletons (I4, J4, K4). The subimages were then colored coded (I5, J5, K5). To visualize mammographic tissue structure, a small RGB image was created where each pixel represents the 360 pixel by 360 pixel subimage that was analyzed (L). A semi-transparent overlay was also constructed to highlight mammographic tissue subtypes on the mammogram (M).

To objectively determine the optimal scale range for fitting power-law curves in *D*(*q, a*) vs. *log*_2_(*a*) and *h*(*q, a*) vs. *log*_2_(*a*) plots, a window was varied along *log*_2_(*a*). The window was defined by a lower bound (*a*_*max*_) and an upper bound (*a*_*min*_) of *a*, varying from *log*_2_*a*_*min*_ = 0, 0.1, …, 2.1 and from *log*_2_*a*_*max*_ = 2.0, 2.1, …, 4.9 respectively, in *σ*_*w*_ units, where *σ*_*w*_ = 7 pixels. All possible combinations of *a*_*min*_ and *a*_*max*_ with a window width being at least *log*_2_*a*_*max*_ – *log*_2_*a*_*min*_ = 1.0 wide, were considered. For each such (*a*_*min*_, *a*_*max*_) window, *h*(*q*) and *D*(*q*) were calculated, along with the goodness of fit *R*^2^ of *h*(*q* = 0), denoted *R*^2^_*h*(*q*=0)_. Additionally, the weighted standard deviation of *h* across all *q* values, denoted *sd*_*w*_, and the weighted average of *R*^2^ of *h*(*q*) over all values of *q*, denoted <*R*^2^_w_>, were also calculated, according to the weights in Marin, et al. [15]. The further consideration of (*a*_*min*_, *a*_*max*_) windows was subject to the fulfillment of several conditions. The first requirement was that the support dimension, represented by *D*(*q* = 0), fell within the range of 1.7 to 2.5, considering the potential impact of finite size effects on the multiplication of maxima lines as the scale parameter *a* approached 0. A window was only considered if it had an *R*^*2*^*h(0)* value exceeding 0.90, ensuring that the *h*(*q* = 0) curve was linear enough to provide a dependable exponent. A low weighted standard deviation for *h*, specifically *sdw* < 0.06, was also essential to exclude subregions demonstrating multifractal scaling. Finally, the condition <*R*^*2*^*w*> > 0.90 was imposed to guarantee that all *h*(*q, a*) curves were sufficiently linear, with greater weight allocated to those closer to *q* = 0.

Based on the resulting *H*, each subregion was classified into one of three groups: fatty tissue (*H* ≤ 0.45, **Fig. 1I5**), active dense tissue (*0.45<H<0.55*, **Fig. 1J5**), or passive dense tissue (*H* ≥ 0.55, **Fig. 1K5**). The area (cm^2^) of each tissue type was estimated for all four mammographic views (**Fig. 1L, 1M**). The right and left MLO and CC views were used to calculate both breasts’ maximum area of passive and active dense tissue to obtain a score for each breast.

### Statistical Methods

The amount of passive and active dense tissue was predicted by creating linear mixed-effects (LME) models. The fixed effects for this model included time (to diagnosis for cancer cases and to the last visit for controls), cancer status, and breast status. An interaction term between time and breast status was fitted to test the hypothesis of changes occurring in dense breast tissue in tumorous breasts vs non-tumorous breasts (**Table 1**). Random effects included breast (left or right) nested within participant nested within case control strata obtained from age-matching. The correlation between repeated visits was modeled using autocorrelation of order 1. All statistical analyses were performed in R [37].

**Table 1:** Description of dependent variables and fixed effects used in the linear mixed effects model.

| Type | Name | Description |
| --- | --- | --- |
| Dependent | Active area | Area of active dense tissue |
|  | Passive area | Area of passive dense tissue |
| Fixed effects | Cancer status | Control: No history of cancer<br>Cancer: Pathology confirmed malignancy detected at last screening exam |
|  | Breast status | No Tumor: Breast does not contain a tumor at last screening exam<br>Tumor: Breast contains a tumor at last screening exam |
|  | Time | Controls: Time since last screening exam<br>Cancer cases: Time since last screening exam before diagnosis |

## RESULTS

Mammogram data and accompanying pathology reports were collected from 50 patients (27 controls and 23 malignant cases), with mammograms obtained using Hologic’s Selena Lorad (Malborough, MA). The patients’ age at the last screening for malignant cases and the time of the last visit for controls ranges from 40 to 85, with a mean age of 65.39 ± 10.03 for malignant cases and 59.56 ± 10.84 for controls. Age-matching with 2 years resulted in keeping 83.7% (24/27 controls and 17/23 malignant cases) of the data and within 5-years, resulted in keeping % (26/27 controls and 20/23 malignant cases) of the data (**Table 2**). Fatty, active, and passive dense tissue were identified in the four bilateral standardized mammographic views for both controls and malignant cases at each time point (**Fig. 2**).

**Table 2:** Cohort description of the primary dataset age-matched within 2 years and the secondary dataset age-matched within 5 years.

| Characteristic | Primary dataset |  | Secondary dataset |  |
| --- | --- | --- | --- | --- |
|  | Cancer, N = 17<sup>†</sup> | Control, N = 24<sup>†</sup> | Cancer, N = 20<sup>†</sup> | Control, N = 26<sup>†</sup> |
| Age | 63.24 (10.30) | 61.00 (10.29) | 65.10 (10.74) | 60.31 (10.31) |
| Number of screening exams |  |  |  |  |
| 4 | 1 (5.9%) | 3 (13%) | 1 (5.0%) | 3 (12%) |
| 5 | 0 (0%) | 4 (17%) | 1 (5.0%) | 4 (15%) |
| 6 | 9 (53%) | 5 (21%) | 9 (45%) | 6 (23%) |
| 7 | 5 (29%) | 11 (46%) | 6 (30%) | 11 (42%) |
| 8 | 2 (12%) | 1 (4.2%) | 3 (15%) | 2 (7.7%) |
| BI-RADS breast density score |  |  |  |  |
| A | 1 (5.9%) | 7 (29%) | 1 (5.0%) | 7 (27%) |
| B | 9 (53%) | 9 (38%) | 12 (60%) | 11 (42%) |
| C | 6 (35%) | 7 (29%) | 6 (30%) | 7 (27%) |
| D | 1 (5.9%) | 1 (4.2%) | 1 (5.0%) | 1 (3.8%) |
| <sup>†</sup> Mean (SD); n (%) |  |  |  |  |

**Figure 2:**
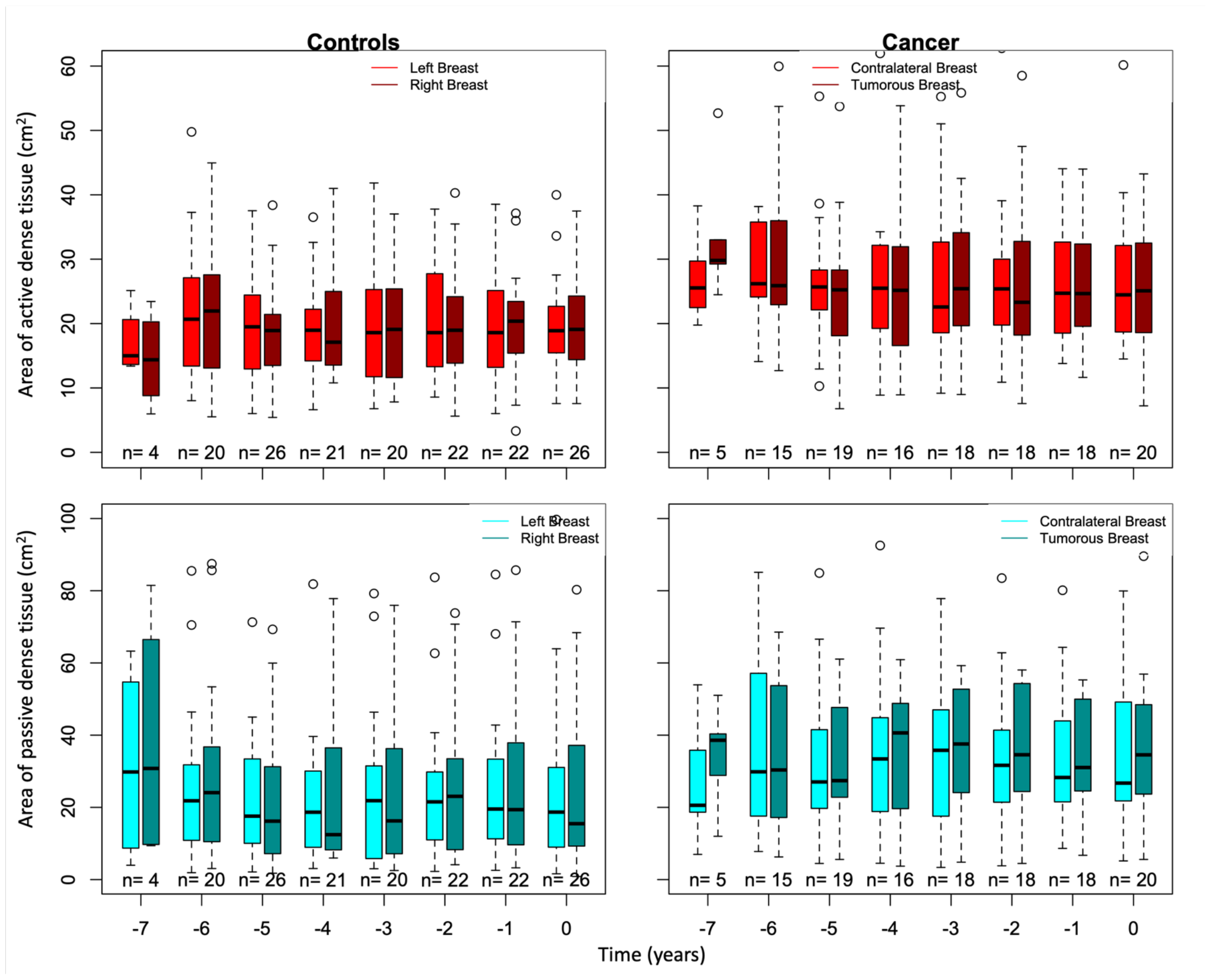
Boxplots of the area of active dense tissue (top row) and passive dense tissue (bottom row) for controls left and right breasts and cancer cases tumorous and contralateral breasts over time using the primary dataset.

Using the primary dataset, which includes controls and cancer cases age-matched within 2 years, the amount of passive and active dense tissue differed between cancer and controls (**Table 3A**). Cancer cases showed 10.30 cm^2^ (CI = 0.75 − 19.85) more passive dense tissue than the average 20.46 cm^2^ found in controls (*p* = 0.036), and 6.34 cm^2^ (, CI = 0.88 − 11.79) more active dense tissue than the 18.55 cm^2^ found in controls (*p* = 0.025). The amount of passive dense tissue also was affected by the breast status (i.e. tumor or no tumor). Breasts that contained a tumor had t 4.16 cm^2^ (CI = 0.61 − 7.70) more passive dense tissue than breasts without tumors (*p* = 0.023). No time effect was detected for the amounts of passive or active dense tissue in the primary dataset.

**Table 3:** Results from linear mixed-effects model using **A)** the primary dataset with patients age-matched within 2 years and **B)** the secondary dataset with patients age-matched within 5 years.

| **A)** |  |  |  |  |  |  |
| --- | --- | --- | --- | --- | --- | --- |
| *Predictors* | **Area of passive dense tissue** |  |  | **Area of active dense tissue** |  |  |
|  | *Estimates* | *CI* | *p* | *Estimates* | *CI* | *p* |
| (Intercept) | 20.46 | 12.24 – 28.69 | **<0.001** | 18.55 | 15.21 – 21.89 | **<0.001** |
| Cancer Status [Cancer] | 10.30 | 0.75 – 19.85 | **0.036** | 6.34 | 0.88 – 11.79 | **0.025** |
| Time | -0.06 | -0.35 – 0.23 | 0.704 | -0.03 | -0.24 – 0.18 | 0.766 |
| Breast Status [Tumor] | 4.16 | 0.61 – 7.70 | **0.023** | -0.62 | -3.02 – 1.77 | 0.601 |
| Time × Breast Status [Tumor] | 0.36 | -0.26 – 0.98 | 0.258 | -0.35 | -0.79 – 0.10 | 0.128 |
| N | 8 Categorical_Time |  |  | 8 Categorical_Time |  |  |
|  | 2 View |  |  | 2 View |  |  |
|  | 41 Patient_ID |  |  | 41 Patient_ID |  |  |
|  | 17 Age_Matching_Subclass |  |  | 17 Age_Matching_Subclass |  |  |
| Observations | 512 |  |  | 512 |  |  |
| Marginal R<sup>2</sup> / Conditional R<sup>2</sup> | 0.540 / NA |  |  | 0.921 / NA |  |  |

| **B)** |  |  |  |  |  |  |
| --- | --- | --- | --- | --- | --- | --- |
| *Predictors* | **Area of passive dense tissue** |  |  | **Area of active dense tissue** |  |  |
|  | *Estimates* | *CI* | *p* | *Estimates* | *CI* | *p* |
| (Intercept) | 21.75 | 13.38 – 30.12 | **<0.001** | 19.27 | 15.29 – 23.24 | **<0.001** |
| Cancer Status [Cancer] | 10.49 | -1.12 – 22.10 | 0.074 | 8.15 | 1.85 – 14.44 | **0.013** |
| Time | -0.40 | -0.69 – -0.10 | **0.009** | -0.09 | -0.29 – 0.12 | 0.405 |
| Breast Status [Tumor] | 5.95 | 2.06 – 9.85 | **0.004** | -0.20 | -2.37 – 1.97 | 0.854 |
| Time × Breast Status [Tumor] | 0.67 | 0.04 – 1.30 | **0.038** | -0.14 | -0.58 – 0.29 | 0.516 |
| N | 8 Categorical_Time |  |  | 8 Categorical_Time |  |  |
|  | 2 View |  |  | 2 View |  |  |
|  | 46 Patient_ID |  |  | 46 Patient_ID |  |  |
|  | 20 Age_Matching_Subclass |  |  | 20 Age_Matching_Subclass |  |  |
| Observations | 580 |  |  | 580 |  |  |
| Marginal R<sup>2</sup> / Conditional R<sup>2</sup> | 0.548 / NA |  |  | 0.947 / NA |  |  |

The model constructed using the secondary dataset, which included patients age-matched within 5 years and increased the power, showed similar estimates for the main effects and interaction terms in the model (**Table 3B**). Like the results using the primary dataset, the model showed that cancer cases had more active dense tissue than controls (*p* = 0.013), with cancer cases having 8.15 cm^2^ (CI = 1.84 − 14.44) than controls and that breast that contained a tumor had more passive dense tissue than breasts that did not contain a tumor (*p* = 0.004). However, the amount of passive dense tissue was only suggestive of differing between cancer cases and controls (*p* = 0.074). In addition, there was a time effect detected for the amount of passive dense tissue, with the amount of passive dense tissue decreasing by 0.40 cm^2^ (CI = −0.69 − −0.10) per year over time for cancer cases and controls (*p* = 0.009). Furthermore, the amount of passive dense tissue in breasts that contained a tumor increased 0.67 cm^2^ (CI = 0.04 − 1.30) per year more than breasts that did not contain a tumor (*p* = 0.038).

## CONCLUSION

Computer-aided detection and technologies that estimate short-term risk have improved the secondary prevention of breast cancer. However, there has been limited research to determine how the changes in breast tissue structure on mammograms connect to the smaller-scale biophysical processes of cancer onset and progression. In prior work, mammographic breast tissue was classified into subtypes (i.e., fatty, passive dense, and active dense) using *H* obtained from the 2D WTMM method and showed that cancer cases had more passive and active dense tissue at the time of diagnosis. Using a LME model with time, cancer status, breast status, and the interaction between time and breast status to predict the amount of passive and active dense tissue has provided insights into a possible new risk factor for breast cancer. Cancer status was a predictor for passive (*p* = 0.036) and active (*p* = 0.025) dense tissue using the primary dataset. However, when increasing the power, cancer status was a predictor for active dense tissue (*p* = 0.013), while breast status (*p* = 0.004), time (*p* = 0.009), and interaction (*p* = 0.038) were predictors for passive dense tissue. This suggests active dense tissue is a risk for cancer and passive dense tissue is an indication of developing cancer.

One limitation of this study was the small sample size, with no information on race or ethnicity. Furthermore, the mammograms used were “FOR PRESENTATION” images from a single vendor. Since algorithms to produce “FOR PRESENTATION” image from “FOR PROCESSING” image vary by vendor, these results must be validated on images obtained from different vendors. In addition to validating our results on a larger and more diverse data set to overcome the limitations discussed, next steps would be to incorporate passive and active dense tissue measurements into a risk model and explore using images obtained from tomosynthesis and “FOR PROCESSING” images.

## Data Availability

The authors will make the data available upon reasonable request.

## DATA AVAILABILITY STATEMENT

The authors will make the data available upon reasonable request.

## AUTHOR CONTRIBUTIONS

Experimental design: KB, AK. WTMM calculations: AK, BW. Radiological breast density assessments: AH, CC. Design and implementation of statistical experiments: CL, KB. Figure and table preparation: KB, AK. Manuscript writing: KB, AK. All authors have read and approved of the manuscript.

## ACKNOWLEDGEMENTS

We are grateful to Drs Anne Breggia, Ivette Emery, and Joe Schulte from MaineHealth for the mammography database, and to CompuMAINE Lab members Arihant Tallapureddy, Melissa Ham, and Sarah Glatter for the manual delineations. We also thank Drs. Karissa Tilbury, Zheng Wei, and David Bradley, for technical discussions.

## REFERENCES

1. Sung, H., J. Ferlay, R.L. Siegel, M. Laversanne, I. Soerjomataram, A. Jemal, and F. Bray, Global Cancer Statistics 2020: GLOBOCAN Estimates of Incidence and Mortality Worldwide for 36 Cancers in 185 Countries. CA Cancer J Clin, 2021. 71(3): p. 209–249.

2. Loomans-Kropp, H.A. and A. Umar, Cancer prevention and screening: the next step in the era of precision medicine. NPJ Precis Oncol, 2019. 3: p. 3.

3. Greene, H., Cancer prevention, screening, and early detection. Advanced Oncology Nursing Certification Review and Resource Manual. 3rd ed. 2016.

4. Siu, A.L. and U.S.P.S.T. Force, Screening for Breast Cancer: U.S. Preventive Services Task Force Recommendation Statement. Ann Intern Med, 2016. 164(4): p. 279–96.

5. American College of Radiology. Breast Cancer Screening For Elevated-Risk Women. [cited 2023 May 30]; Available from: https://www.acr.org/-/media/ACR/Files/Breast-Imaging-Resources/Care-Toolkit/Provider-Breast-Cancer-Risk-Assessment-Handout.pdf.

6. Ebell, M.H., T.N. Thai, and K.J. Royalty, Cancer screening recommendations: an international comparison of high income countries. Public Health Rev, 2018. 39: p. 7.

7. Mullen, L.A., B. Panigrahi, J. Hollada, B. Panigrahi, E.T. Falomo, and S.C. Harvey, Strategies for Decreasing Screening Mammography Recall Rates While Maintaining Performance Metrics. Acad Radiol, 2017. 24(12): p. 1556–1560.

8. Freer, P.E., Mammographic breast density: impact on breast cancer risk and implications for screening. Radiographics, 2015. 35(2): p. 302–15.

9. Islami, F., A. Goding Sauer, K.D. Miller, R.L. Siegel, S.A. Fedewa, E.J. Jacobs, M.L. McCullough, A.V. Patel, J. Ma, I. Soerjomataram, W.D. Flanders, O.W. Brawley, S.M. Gapstur, and A. Jemal, Proportion and number of cancer cases and deaths attributable to potentially modifiable risk factors in the United States. CA Cancer J Clin, 2018. 68(1): p. 31–54.

10. Guerrero, V.G., A.F. Baez, C.G. Cofre Gonzalez, and C.G. Mino Gonzalez, Monitoring modifiable risk factors for breast cancer: an obligation for health professionals. Rev Panam Salud Publica, 2017. 41: p. e80.

11. Koh, B., D.J.H. Tan, C.H. Ng, C.E. Fu, W.H. Lim, R.W. Zeng, J.N. Yong, J.H. Koh, N. Syn, W. Meng, K. Wijarnpreecha, K. Liu, C.S. Chong, M. Muthiah, H.N. Luu, A. Vogel, S. Singh, K.G. Yeoh, R. Loomba, and D.Q. Huang, Patterns in Cancer Incidence Among People Younger Than 50 Years in the US, 2010 to 2019. JAMA Netw Open, 2023. 6(8): p. e2328171.

12. Zhao, J., L. Xu, J. Sun, M. Song, L. Wang, S. Yuan, Y. Zhu, Z. Wan, S. Larsson, K. Tsilidis, M. Dunlop, H. Campbell, I. Rudan, P. Song, E. Theodoratou, K. Ding, and X. Li, Global trends in incidence, death, burden and risk factors of early-onset cancer from 1990 to 2019. BMJ Oncology, 2023. 2(1): p. e000049.

13. American Cancer Society. Genetic Counseling and Testing for Breast Cancer Risk. [cited 2023 May 18]; Available from: https://www.cancer.org/cancer/types/breast-cancer/risk-and-prevention/genetic-testing.html.

14. Vianna, F.S.L., J. Giacomazzi, C.B. Oliveira Netto, L.N. Nunes, M. Caleffi, P. Ashton-Prolla, and S.A. Camey, Performance of the Gail and Tyrer-Cuzick breast cancer risk assessment models in women screened in a primary care setting with the FHS-7 questionnaire. Genet Mol Biol, 2019. 42(1 suppl 1): p. 232–237.

15. Wengert, G.J., T.H. Helbich, D. Leithner, E.A. Morris, P.A.T. Baltzer, and K. Pinker, Multimodality Imaging of Breast Parenchymal Density and Correlation with Risk Assessment. Curr Breast Cancer Rep, 2019. 11(1): p. 23–33.

16. Gail, M.H. and R.M. Pfeiffer, Breast Cancer Risk Model Requirements for Counseling, Prevention, and Screening. J Natl Cancer Inst, 2018. 110(9): p. 994–1002.

17. World Health Organization. Breast cancer. [cited 2023 May 30]; Available from: https://www.who.int/news-room/fact-sheets/detail/breast-cancer.

18. Arasu, V.A., L.A. Habel, N.S. Achacoso, D.S.M. Buist, J.B. Cord, L.J. Esserman, N.M. Hylton, M.M. Glymour, J. Kornak, L.H. Kushi, D.A. Lewis, V.X. Liu, C.M. Lydon, D.L. Miglioretti, D.A. Navarro, A. Pu, L. Shen, W. Sieh, H.C. Yoon, and C. Lee, Comparison of Mammography AI Algorithms with a Clinical Risk Model for 5-year Breast Cancer Risk Prediction: An Observational Study. Radiology, 2023. 307(5): p. e222733.

19. Acciavatti, R.J., S.H. Lee, B. Reig, L. Moy, E.F. Conant, D. Kontos, and W.K. Moon, Beyond Breast Density: Risk Measures for Breast Cancer in Multiple Imaging Modalities. Radiology, 2023. 306(3): p. e222575.

20. de Vries, C.F., S.J. Colosimo, R.T. Staff, J.A. Dymiter, J. Yearsley, D. Dinneen, M. Boyle, D.J. Harrison, L.A. Anderson, G. Lip, and C.R.C. i, Impact of Different Mammography Systems on Artificial Intelligence Performance in Breast Cancer Screening. Radiol Artif Intell, 2023. 5(3): p. e220146.

21. Bissell, M.J. and W.C. Hines, Why don’t we get more cancer? A proposed role of the microenvironment in restraining cancer progression. Nat Med, 2011. 17(3): p. 320–9.

22. Kenny, P.A., G.Y. Lee, and M.J. Bissell, Targeting the tumor microenvironment. Front Biosci, 2007. 12: p. 3468–74.

23. Ronnov-Jessen, L. and M.J. Bissell, Breast cancer by proxy: can the microenvironment be both the cause and consequence? Trends Mol Med, 2009. 15(1): p. 5–13.

24. Tanner, K., H. Mori, R. Mroue, A. Bruni-Cardoso, and M.J. Bissell, Coherent angular motion in the establishment of multicellular architecture of glandular tissues. Proc Natl Acad Sci USA, 2012. 109(6): p. 1973–8.

25. Strandberg, R., K. Czene, M. Eriksson, P. Hall, and K. Humphreys, Estimating Distributions of Breast Cancer Onset and Growth in a Swedish Mammography Screening Cohort. Cancer Epidemiol Biomarkers Prev, 2022. 31(3): p. 569–577.

26. Batchelder, K.A., A.B. Tanenbaum, S. Albert, L. Guimond, P. Kestener, A. Arneodo, and A. Khalil, Wavelet-based 3D reconstruction of microcalcification clusters from two mammographic views: New evidence that fractal tumors are malignant and Euclidean tumors are benign. PLoS One, 2014. 9(9): p. e107580.

27. Kestener, P., J.M. Lina, P. St-Jean, and A. Arneodo, Wavelet-based multifractal formalism to assist in diagnosis in digitized mammograms. Image Anal Stereol, 2001. 20: p. 169–74.

28. Khalil, A., G. Joncas, F. Nekka, P. Kestener, and A. Arneodo, Morphological analysis of HI features. II. Wavelet-based multifractal formalism. Astrophys J Suppl S, 2006. 165: p. 512–50.

29. Khalil, A., C. Aponte, R. Zhang, T. Davisson, I. Dickey, D. Engelman, M. Hawkins, and M. Mason, Image analysis of soft-tissue in-growth and attachment into highly porous alumina ceramic foam metals. Med Eng Phys, 2009. 31(7): p. 775–83.

30. Arneodo, A., N. Decoster, and S. Roux, A wavelet-based method for multifractal image analysis. I. Methodology and test applications on isotropic and anisotropic random rough surfaces. Eur Phys J B, 2000. 15: p. 567–600.

31. Decoster, N., A. Arneodo, and S. Roux, A wavelet-based method for multifractal image analysis. II. Applications to synthetic multifractal rough surfaces. Eur Phys J B, 2000. 15: p. 739–64.

32. Marin, Z., K.A. Batchelder, B.C. Toner, L. Guimond, E. Gerasimova-Chechkina, A.R. Harrow, A. Arneodo, and A. Khalil, Mammographic evidence of microenvironment changes in tumorous breasts. Med Phys, 2017. 44(4): p. 1324–36.

33. Gerasimova-Chechkina, E., B.C. Toner, K.A. Batchelder, B. White, G. Freynd, I. Antipev, A. Arneodo, and A. Khalil, Loss of Mammographic Tissue Homeostasis in Invasive Lobular and Ductal Breast Carcinomas vs. Benign Lesions. Frontiers in Physiology, 2021. 12(595).

34. American College of Radiology, B.I.R.C., ACR BI-RADS atlas : breast imaging reporting and data system. 5th ed. Breast imaging reporting and data system. 2013, Reston, VA: American College of Radiology.

35. Ho, D.E., Imai, K., King, G., Stuart, E.A., MatchIt: Nonparametric Preprocessing for Parametric Causal Inference. Journal of Statistical Software, 2011. 42(8): p. 1–28.

36. Schneider, C.A., Rasband, W.S., Eliceiri, K.W., NIH Image to ImageJ: 25 years of image analysis. Nature Methods, 2012. 9: p. 671–675.

37. R Core Team. R: A language and environment for statistical computing. R Foundation for Statistical Computing, Vienna, Austria. 2021; Available from: https://www.R-project.org/.

